# Antigen-experienced CXCR5^-^ CD19^low^ B cells are plasmablast precursors expanded in SLE

**DOI:** 10.1101/2021.05.25.21257784

**Authors:** Franziska Szelinski, Ana Luisa Stefanski, Annika Wiedemann, Eva Schrezenmeier, Hector Rincon-Arevalo, Karin Reiter, Marie Lettau, Van Duc Dang, Sebastian Fuchs, Andreas P. Frei, Tobias Alexander, Andreia C. Lino, Thomas Dörner

## Abstract

B cells play a critical role in the pathogenesis of systemic lupus erythematosus (SLE). We analysed two independent cohorts of healthy donors and SLE patients using a combined approach of flow and mass cytometry. We have found that IgD^-^ CD27^+^ switched and atypical IgD^-^CD27^-^ memory B cells, which are increased in SLE, represent heterogeneous populations composed each of three different subsets, such as CXCR5^+^CD19^int^, CXCR5^-^CD19^high^ and CXCR5^-^CD19^low^. Here, we characterize a hitherto unknown antigen-experienced CXCR5^-^CD19^low^ B cell subsets enhanced in SLE and carrying a plasmablast (PB) phenotype enriched for switched immunoglobulins, and expressing CD38, CD95, CD71, *PRDM1, XBP-1*, and *IRF4*. CXCR5^-^CD19^low^ resemble activated B cells with a characteristically diminished B cell receptor responsiveness. CXCR5^-^CD19^low^ B cells increased with PB frequencies in SLE and upon BNT162b2 vaccination suggesting their interrelationship. Our data suggest that CXCR5^-^CD19^low^ B cells are precursors of plasmablasts, thus co-targeting this subset may have therapeutic value in SLE.

## Introduction

Various abnormalities of the B cell lineage have been identified in systemic lupus erythematosus (SLE), a chronic autoimmune disease characterized by autoantibody production and pathogenic autoimmune complex formation. Abnormalities include increased peripheral plasmablasts (PB)^1^, including expanded IgG and IgA producers^2^ as well as altered composition of the B cell compartment with increased frequencies of IgD^-^CD27^+^switched and atypical IgD^-^CD27^-^ memory B cells. How these observation are related remains unclear. Increased switched memory or IgD^-^CD27^-^ B cells were found in rheumatic diseases such as SLE^3^ and RA^4, 5, 6^ as well as in peripheral blood or tissue in various inflammatory diseases, such as chronic inflammatory bowel^7^ or Alzheimer’s disease^8^.

In SLE, expansion of switched memory and IgD^-^CD27^-^, (also called double negative, DN) B cells, are features of patients with chronic inflammatory diseases. This shift in the B cell distribution is not seen in new onset patients even though their B cell distribution also differs from healthy donors (HD)^9^. These observations emphasize that enlargement of switched memory and DN B cells are an important characteristic of chronic inflammation^9^. In lupus nephritis (LN), a severe complication of SLE, DN B cells correlated with 24-h urine protein excretion and inversely with glomerular filtration rate^10^. Interestingly, this population was found diminished in LN patients during remission suggesting their potential pathogenic involvement and raising the possibility that DN B cells can serve as a prognostic biomarker in lupus nephritis^10^.

More recently, it became evident that DN B cells represent a heterogeneous subset, including age-/autoimmune-associated B cells (ABCs)^11, 12, 13^, Syk^++^ B cells^14^ and double negative 2 (DN2: IgD^-^CD27^-^CXCR5^-^CD11c^+^) populations^15^. All these subsets share partially overlapping characteristics and are linked to disease activity and autoantibody formation. Besides these alterations of B cell subsets residing among DN B cells, impaired chemokine receptor expression^16^ and reduced responsiveness upon BCR stimulation^17, 18, 19^ have been reported for these B cells in patients with SLE suggesting their distinct involvement in chronic autoimmunity.

This study further delineates the heterogeneity of switched memory (mem) and DN memory B cells in healthy donors and SLE patients. Using a combined flow and mass cytometry approach, we identified an enhanced CXCR5^-^CD19^low^ B cell subset in the switched memory and DN compartments (mem^low^ and DN^low^) in SLE carrying distinct functional characteristics of antigen-experienced B cells distinct from previously known CXCR5^+^CD19^int^ (mem^int^ and DN^int^) and CXCR5^-^CD19^high^ (mem^high^ and DN^high^) B cells which showed a close relationship with peripheral PB. Our data provide multiple new insights, including B cell differentiation abnormalities in SLE relevant for therapeutic considerations targeting B cells in SLE.

## Material and methods

### Patients

Peripheral blood was obtained from 79 SLE patients meeting the SLICC criteria and from 70 healthy individuals. All study participants gave written consent to participate in this cross-sectional study which was approved by the ethics’ committee of the Charité Berlin.

### Antibodies

See Supplementary table 2 for antibodies used for flow cytometry analysis and Supplementary table 3 for antibodies used for mass cytometry analysis.

### Whole blood stainings

Erythrolysis of EDTA anticoagulated blood was performed according to manufacturer’s protocol using Pharm Lyse (BD). FcR blocking reagent (Miltenyi Biotec) was added to cell suspension before cells were stained for CD3, CD14, CD20, CD19, CD27, IgD, CD38, CD95 and CXCR5. Frequencies of cells were determined using a BD Canto II cytometer.

### Peripheral blood mononuclear cells (PBMCs) isolation and cryopreservation

PBMCs were isolated from EDTA anticoagulated whole blood for flow cytometry stainings and stimulation experiments. Li-Hep-anticoagulated blood was used for CyTOF analysis. Therefore, whole blood was diluted in PBS (Biochrom), layered over Ficoll-Paque PLUS (GE Healthcare Bio-Sciences) and centrifuged. PBMCs were harvested and washed twice with PBS before cell counting. PBMCs were processed immediately or cryopreserved for CyTOF analysis. For this, up to 10×10^6^ PBMCs were diluted in 1:10 DMSO in FBS and cooled down to -80°C using CoolCell Cell Freezing Container (Biocision) before storing at -80°C until further analysis.

### Viability assay for discrimination of live and dead cells

PBMCs were labelled with Blue fluorescent reactive dye (Molecular Probes Invitrogen) 1:1000 in PBS according to the manufacturer’s recommendation and washed with PBE. Live/dead stained PBMCs were directly suspended in cold MACS rinsing buffer (with BSA; Miltenyi Biotec) for phenotyping or in pre-warmed RPMI 1640 (with GlutaMAX, Life Technologies) for BCR stimulation assays.

### Backbone surface staining of PBMCS for subset identification and phenotyping

Viability labelled cells were pre-treated with FcR blocking reagent (Miltenyi Biotec) for 5 min before being stained with anti-CD19, CD27, CD38, IgD, CD14, CD13, CD10, CD24, CD11c and CD95 as backbone staining and for phenotyping in addition with anti-CD71, -PD1 and -PD-L1. Median fluorescence intensities (MFIs) or frequencies of positive cells were determined using BD LSR Fortessa x-20 (Beckton Dickinson).

### Heat map of foldchange

To evaluate expression of surface marker of subsets in comparison to expression levels on the main B cell populations, log_2_(fold changes) of median FI or frequencies from the flow cytometry data were plotted as heat map.

### Staining for immunoglobulin isotypes

Viability labelled cells were stained with anti-CD19, CD20, CD27, CD38, CD14, CD13, CD95, CXCR5, IgD, IgM, IgG and IgA. Data was optained using BD LSR Fortessa x-20 (Beckton Dickinson). Frequencies of cells positive for the different isotypes were collected.

### High-dimensional single-cell proteomics analysis using CyTOF

Frozen PBMCs were thawed and resuspended at 37°C in 1:10 FCS in IMDM. Further sample processing was done as previously described^20^. Samples where measured using a Helios mass cytometry instrument (Fluidigm).

### Vaccination of healthy individuals with BNT162b2

B cell subsets of healthy individuals were analyzed before administration, 7, 14, 21 days after first vaccination and 7 days after boost with BNT162b2 (Comirnaty®).

### Data analysis and UMAP plotting

Flow and mass cytometry data were analyzed using FlowJo (version 10.6.1, TreeStar). Flow cytometry data was pre-gated on IgD^-^ B cells, down sampled to 59.000 events each per cohort of HD or SLE, respectively and clustered by CXCR5, CD19, CD38, CD27, CD10, CD71, CD95 and IgD using dimension reduction algorithm UMAP^21^ plugin in FlowJo. Mass cytometry data was pre-gated on IgD^-^ B cells excluding plasmablasts, down sampled to 8841 cells and clustered by CXCR5, CD19, CD38, CD27, CD11c and IgM using UMAP plugin. As settings for both UMAPs we selected the Euclidean distance function, nearest neighbour value of 15 and a minimum distance of 0.5.

### BCR stimulation for analysis of intracellular phosphorylation

Cells (3× 10^6^) stained for viability, blocked and stained with modified backbone surface staining (excluding CD24, CXCR5 but including CD22) and equilibrated for 1h at 37°C in RPMI before stimulation with IgG/IgA/IgM (H+L) F(ab’)_2_ (15 µg/ml) for 5, 8 or 15 min. For baseline, control cells were incubated for 5 min with RPMI. Adding pre-warmed Lyse/Fix buffer (BD) at the respective time-points stopped the stimulation. After washing Phosflow Perm II Buffer (BD) was added to permeabilize cells overnight at - 20°C. Cells were then stained for CD24, CXCR5 and the intracellular targets Syk and pSyk (Y352). To investigate BCR response and phosphorylation kinetics, median FIs of Syk, pSyk (Y352) and CD22 were determined.

### Gene expression analysis of B cell subsets

Isolated PBMCs were enriched for B cells by depleting CD3, CD14 and CD235a via MACS microbeads (Miltenyi Biotec) according to the manufacturer’s instruction. Cells were stained with CD19, CD20, CD27, CD38, CD3, CD14, CXCR5 and IgD for sorting. Naïve, pre-switched, total memory and PBs as well as DN^int^, DN^low^ and DN^high^ subsets were sorted and purity check was performed using Sony Sorter MA900. Cells were counted using MACSquant (Miltenyi Biotec). Cell suspension were spin down, resuspended in HTG lysis buffer at a concentration of 200 cells/µl and stored at -80 °C until further processing by HTG (Tucson, AZ). Samples with less than 28% of counts allocated to positive controls probes, read depth above 750,000 and expression variability above 0,094 were analyzed.

### Statistical evaluation

Data analysis was performed using GraphPad Prism® (Version 9). Statistical significance was considered for p values less than 0.05 and depicted as follows: \**p* ≤ 0.05, \*\**p* ≤ 0.01, \*\*\**p* ≤ 0.001, \*\*\*\**p* ≤ 0.0001. Statistical significance between HD and SLE was determined using two-tailed Mann-Whitney-U (MWU) test. To determine statistical significance between the three populations Kruskal-Wallis-Test (KWT) followed by the Dunn’s multiple comparison test was performed. Unless stated otherwise, scatter and bar plots represent means ± SD and Box-Whisker plots show median and range.

## Results

### CD19^low^ memory and double negative B cells are increased in SLE patients

We and others have reported several abnormalities within the B cell compartment in SLE patients suggesting their pathogenic relevance. In this regard, IgD^-^CD27^-^ double negative (DN) B cells especially those co-expressing CD95^22^ and IgD^-^CD27^+^ switched memory B cells are increased in SLE^1, 3^. Therefore, we analyzed these B cell subsets in further detail (Supplementary Fig. S1) and found that switched memory B cells and DN B cells show a similar pattern regarding expression of CXCR5 and CD19, thus, independent of their CD27 expression. Notably, the expression of these molecules subdivided both compartments into three populations (Fig. 1A, Supplementary Fig. S1A). CXCR5^+^CD19^int^ (mem/DN^int^) and CXCR5^-^ CD19^high^ (mem/DN^high^) B cells have been previously identified and described in HD and SLE with increased frequencies of the CXCR5^-^CD19^high^ fraction in SLE^15^. Here, we identified a novel B cell subset within both switched memory B cells and DN B cells that is CXCR5^-^CD19^low^ (mem/DN^low^).

**Figure 1:**
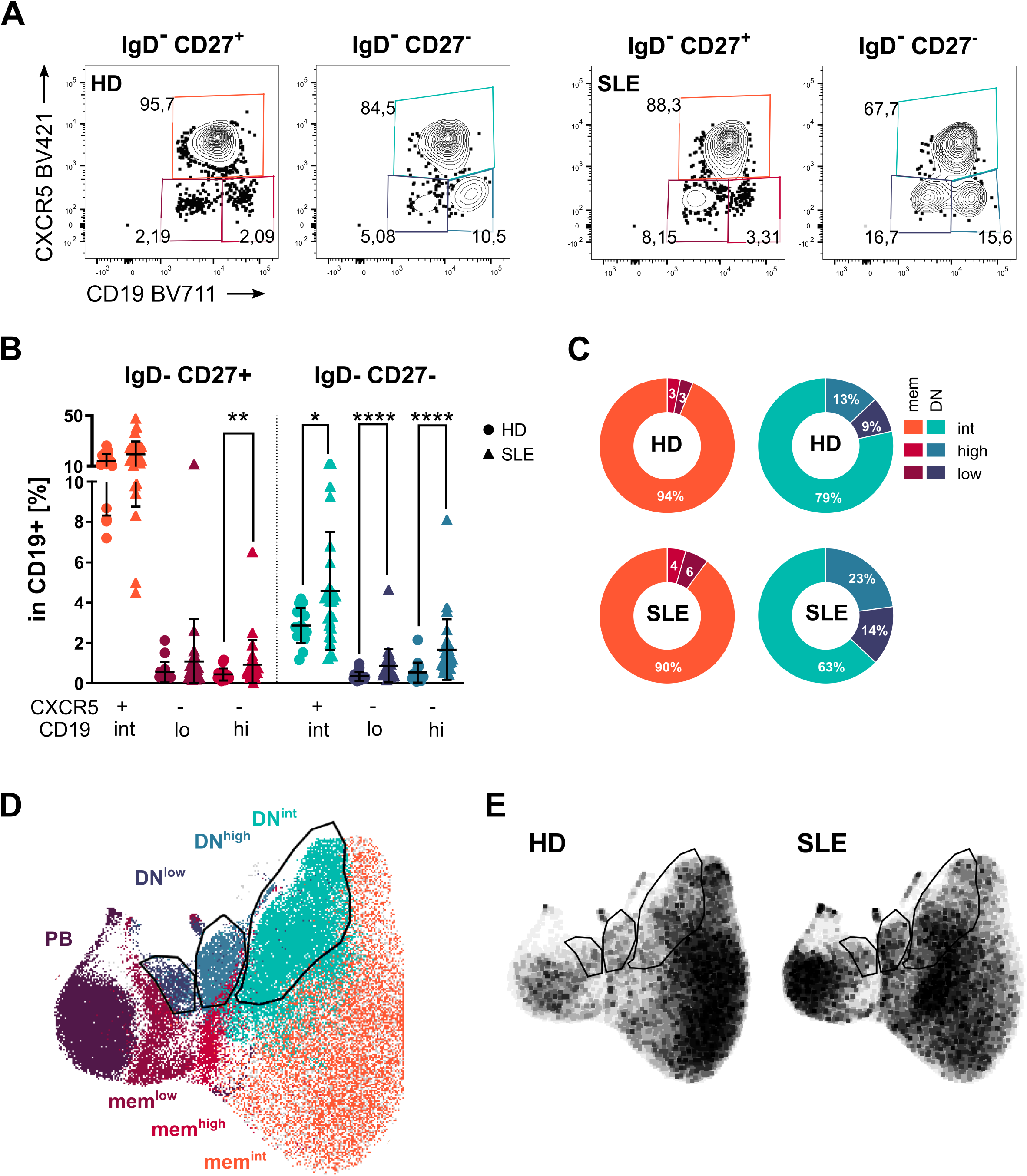
CD19^low^CXCR5^-^ B cells are increased in SLE patients. **(A)** Representative flow cytometry contour plots of one healthy donor and one SLE patient showing CD19 and CXCR5 expression in IgD^-^CD27^+^ or IgD^-^CD27^-^ B cells. **(B)** Distribution of CD19^+^ B cell frequencies of the six populations CXCR5^+^CD19^int^ (mem^int^/DN^int^), CXCR5^-^CD19^low^ (mem^low^/DN^low^), CXCR5^-^CD19^high^ (mem^high^/DN^high^) of IgD^-^CD27^+^ memory or IgD^-^CD27^-^ DN B cells respectively for HD (n=16, dots) and patients with SLE (n=28, triangles). **(C)** Distribution of subsets within switched memory (IgD^-^CD27^+^) and DN (IgD^-^CD27^-^) in HD (n=16) and SLE (n=28) **(D)** Overlay of UMAP clustering of pre-gated IgD^-^ B cells (59000 events each per cohort of HD or SLE, respectively) and manual gated IgD^-^CD27^-^ subsets. Gates of DN^int^, DN^low^ and DN^high^ populations are indicated. **(E)** Comparison of UMAP density plots clustering corresponding B cell subsets of HD and SLE. (MWU between selected relevant comparisons:\**p* ≤ 0.05, \*\**p* ≤ 0.01, \*\*\**p* ≤ 0.001, \*\*\*\**p* ≤ 0.0001)

We found that DN^low^ B cells are significantly increased among CD19^+^ B cells in SLE compared to HD (Fig. 1A). While only mem^high^ B cells were increased in the switched memory compartment, all of the three subsets within the DN fraction were significantly increased in SLE patients (Fig. 1B). DN B cells were enriched for CXCR5 negative CD19^low^ and CD19^high^ B cell subsets compared to the CD27^+^ (mem) compartment and in general remarkably expanded in SLE (Fig. 1 C). Next, we applied the dimension reduction algorithm UMAP ^21^ to cluster IgD^-^ B cells. Using this approach, we identified mem/DN CD19^int^, mem/DN^high^ and mem/DN^low^ as distinct populations (Fig. 1D). In this UMAP, both CD19^low^ populations clustered together with CD27^++^CD38^++^ PB (Fig. 1D). Comparison of the clusters obtained from HD with SLE patients revealed an increased density of the corresponding subsets in SLE (Fig. 1E), consistent with their significant expansion in this condition.

### CD19^low^ B cell subsets display a plasmablast-like phenotype

Next, we characterized DN^int^, DN^high^ and DN^ow^ B cell subsets for the expression of several surface molecules including lineage, differentiation and activation markers. The resulting expression patterns of CD27, CD19, CXCR5, CD24, CD71, CD95, CD38 and CD11c is visualized by colour code in the dimension reduced UMAP (Fig. 2A). With this analysis, we found that the subsets defined by their distinct CD19 and CXCR5 expression allowed further differentiation by different expression profiles of CD24, CD71, CD95, CD38 and CD11c among the IgD^-^CD27^-^ subsets (Fig. 2A).

**Figure 2:**
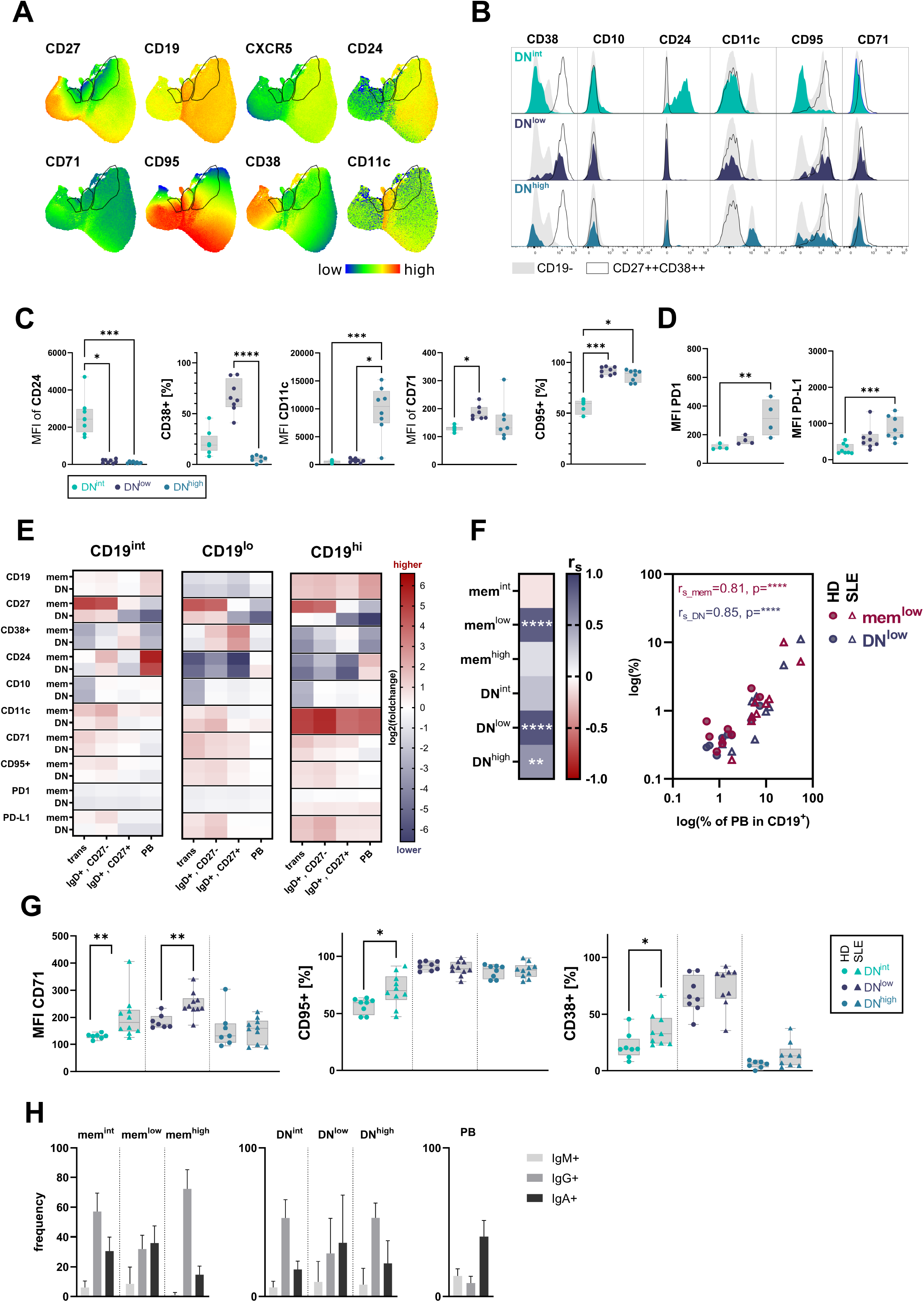
Expression patterns of surface markers differ between DN^low^, DN^high^ and DN^int^ subsets but are comparable to corresponding memory subsets. **(A)** Colour coded mean signal intensity of CD27, CD19, CXCR5, CD24, CD71, CD95, CD38 and CD11c presented as UMAP. Gates indicate the clustered areas of DN^low^ (left gate), DN^high^ (middle gate) and DN^int^ (right gate). **(B)** Representative histograms of flow cytometry data of DN subsets of one healthy donor showing surface marker expression in comparison to CD19^-^ cells (grey) and CD27^++^CD38^++^ plasmablasts (black line). **(C)** Box and whisker plots of median FI of CD24, CD11c, CD71 and frequencies of CD38^+^ or CD95^+^ of HD (n=8) for each subset of DN **(D)** and median FIs of PD1 and PD-L1 in HD (n=8).(test: MWU) **(E)** Heat map showing log_2_(fold change) of median FI of surface marker expression by mem/DN^int^, mem/DN^low^ and mem/DN^high^ subsets related to main B cell populations of transitional, naïve (IgD^+^CD27^-^), pre-switched (IgD^+^CD27^+^) and plasmablast (PB). Red indicating markers that are higher expressed on the subsets of interest and blue indicating a lower expression of the marker on the subsets compared to the main B cell populations. **(F)** Heatmap of Spearman correlation coefficient (r_s_) and scatterplot of correlation between PBs and mem/DN^int^, mem/DN^low^ and mem/DN^high^ subset frequencies of HD (n=8) and SLE (n=10). **(G)** Box and whisker plots of median FI of CD71, CD24, CD11c and frequencies of CD38+ or CD95+ of SLE (triangles, n=10) and HD (dots, n=8) for each DN subset.(test: KWT) **(H)** Frequencies of immunoglobulin isotypes per DN and mem subset, respectively of HD (n=6). (Significance levels: \**p* ≤ 0.05, \*\**p* ≤ 0.01, \*\*\**p* ≤ 0.001, \*\*\*\**p* ≤ 0.0001)

We found that CD24, a marker with a dynamic expression pattern throughout B cell maturation and absent in antibody producing cells^23^, was not present in both subsets with low CD19 and lacking CXCR5 expression (mem/DN^low^) in contrast to the DN^int^ population.

Besides CD19 expression the main discriminators between the DN^low^ and DN^high^ populations were CD38 and CD11c (Fig. 2A, B). As previously described, DN^high^ are CD11c^high^ but lack CD38 expression^15, 20^. In contrast, the majority of DN^low^ B cells express CD38 (Fig. 2B, C and Supplementary Fig. 1B). Expression of CD71, a marker for early B cell activation^24^, was upregulated on the surface of CXCR5^-^ CD19^low^ B cells, comparable to levels found on plasmablasts (Fig. 2C). CD95 was expressed by the majority of DN^low^ and DN^high^ cells but not on DN^int^.

Subsequently, we evaluated the expression of inhibitory receptor PD-1 that is upregulated on B cells upon activation^19, 25^ and its ligand PD-L1. Surface expression of PD-L1 and PD-1 was only found enhanced in DN^high^ (Fig. 2D, Supplementary Fig. 1C).

Most noteworthy, each of the subsets of DN differed from the corresponding subsets in the switched memory compartment mainly by the expression of CD27 while the other makers were expressed in a similar pattern (Fig. 2E, Supplementary Fig. 1). This, suggesting that memory and DN population consist of complementary subsets.

Comparison of the newly identified subsets with conventional transitional (CD10^+^CD24^+^CD38^+^), naïve (IgD^+^CD27^-^) or pre-switched memory (IgD^+^CD27^+^) B cells and PB (CD27^++^CD38^++^) demonstrated that mem/DN subsets are similar to PB, expressing comparable levels of CD19, CD24, CD10, CD11c, CD71, PD1, PD-L1 and CD95. Main differences between PB and mem/DN^low^ cells were a diminished/absent expression of CD27 and a slightly lower CD38 expression among both mem^low^ and DN^low^ B cell subsets (Fig. 2E). It needs emphasis that the frequencies of mem/DN^low^ B cell strongly correlated with those of PB (Fig. 2F) in SLE patients as well as healthy controls further supporting their potential relationship.

Since we saw differences in the frequencies we also checked for qualitative differences between HD and SLE. When comparing expression profiles of the three DN B cell subsets between HD and SLE patients, we found that expression levels of proliferation marker CD71 and frequencies of activation markers CD95^+^ and CD38^+^ cells were increased within the DN^int^ (Fig. 2G) but not the mem^int^ (Supplementary Fig.1D) population of SLE patients. The expression of CD71 was enhanced in both DN^low^ and mem^low^ in SLE (Fig. 2G, Supplementary Fig. 1D). Regarding Ig isotype expression, we found that mem/DN^int^ and mem/DN^high^ mainly express IgG while mem/DN^low^ express IgG and IgA to a similar extend. (Fig. 2H).

### CD19^low^ B cells are characterized by a distinct activation and differentiation profile

Next, we validated our findings in an independent cohort of 27 SLE patients and 18 healthy donors using CyTOF including expression of activation markers and checkpoint molecules (Fig. 3, Supplementary Fig. 2). Initially, we confirmed the increased frequency of all three subsets within the DN compartment in SLE patients as shown by the UMAP plot by increased DN^int^, DN^high^ and DN^low^ clusters (Fig. 3A). Besides the significantly increased DN subsets in patients, mem^low^ B cells were also substantially increased in SLE (Fig. 3B), corroborating the flow cytometry findings noted above.

**Figure 3:**
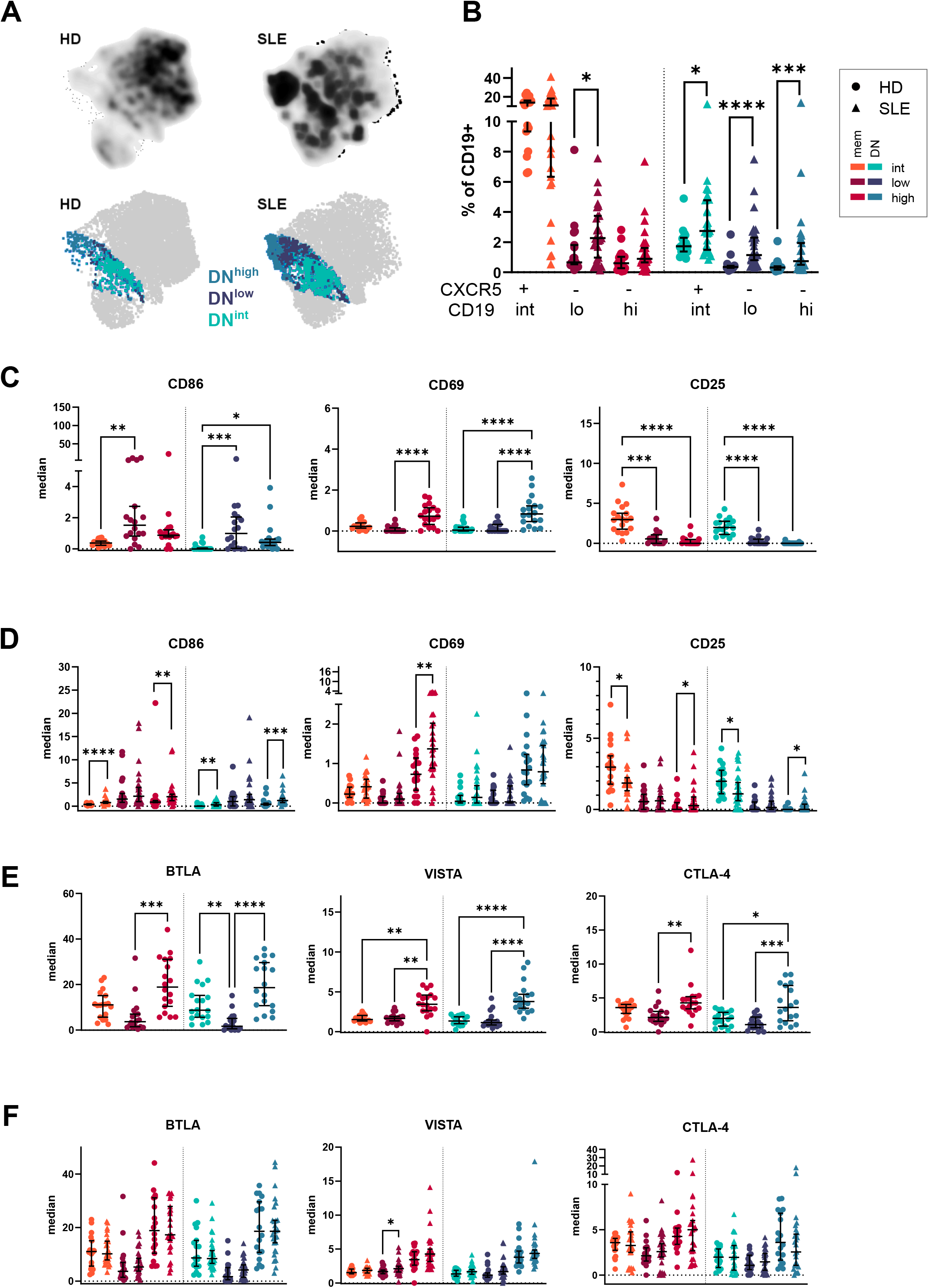
CXCR5^-^CD19^low^ populations show an activated phenotype. **(A)** CyTOF derived UMAP plots (showing 8841 events per cohort of HD and SLE) of IgD^-^ B cells. Top row shows density plots of HD and SLE. Bottom row shows distribution of gated for DN^int^, DN^low^ and DN^high^. **(B)** Frequencies among total CD19^+^ B cells of mem/DN^int^, mem/DN^low^ and mem/DN^high^ of HD (dots, n=18) and patients with SLE (triangles, n=24). Expression of activation markers CD86, CD69 and CD25 by mem/DN^int^, mem/DN^low^ and mem/DN^high^ for **(C)** comparison of subsets (test: MWU) or **(D)** HD vs SLE (test: KWT). Scatter plots show the expression of immune checkpoint molecules for comparison **(E)** in between subsets or **(F)** between HD and SLE. (Data is shown as median + 95%CI of [n(HD/SLE) = 18/24]. (Significance levels: \**p* ≤ 0.05, \*\**p* ≤ 0.01, \*\*\**p* ≤ 0.001, \*\*\*\**p* ≤ 0.0001)

Expression of early activation markers such as CD86, CD69 and CD25, both mem/DN^low^ and mem/DN^high^ were indicative of an activated phenotype with increased CD86 and in case of mem/DN^high^ also increased CD69 expression compared to mem/DN^int^. In contrast, mem/DN^high^ and mem/DN^low^ populations expressed less CD25 compared to mem/DN^int^ (Fig. 3C).

SLE patients expressed more CD86, CD69, and CD25 on the surface of mem^high^ B cells and were characterized by elevated CD86 and CD25 expression on DN^high^ compared to HD. In general, mem/DN^int^ cells of patients with SLE expressed more CD86 and diminished CD25 surface expression (Fig. 3D) indicating their increased activation status.

Additionally, we determined CD45RA and CD45RO expression, known to be differentially expressed throughout B cell differentiation^26, 27^. CD45RA was increased on switched memory B cells compared to DN, but its expression was comparable among the three subsets analyzed (Supplementary Fig. 2 C). When comparing the expression in SLE, we found that CD45RA expression was reduced on DN^high^ cells from patients in comparison with HD (Supplementary Fig. 2 D).

Co-stimulatory and co-inhibitory immune checkpoints (CPMs) regulate and modulate immune cells and play an important role in fine tuning the immune response^20, 28, 29, 30, 31^. Therefore, we analyzed the expression profiles of various CPMs among the subsets of interest. Of particular note, mem/DN^high^ upregulated immune checkpoint molecules, such as BTLA, VISTA and CTLA-4 (Fig. 3E, Supplementary Fig. 2 E, F) suggesting a shared functionality. While VISTA and CTLA-4 expression by mem/DN^low^ cells was comparable to the levels found on the mem/DN^int^, BTLA was downregulated (Fig.3E).

SLE patients expressed higher levels of VISTA on mem^low^ B cells (Fig. 3F) but otherwise did not differ from findings in HD. Overall, CD19^low^ B cells showed a strikingly reduced expression of checkpoint molecules independent of their CD27 expression.

### CD19^low^ subsets correlate with plasmablasts generated in HD upon vaccination with BNT162b2

Since mem^low^ and DN^low^ are increased in SLE and correlated with PB, we asked whether an acute immune response resulting in PB formation would be accompanied by alterations and expansion of those subsets in HD. Therefore, we monitored B cell subsets in HD on day 0, 7, 14 and 21 after single-dose administration of BNT162b2 vaccine and 7 days after boost (Fig. 4A). Of particular note, this vaccine is able to elicit a striking T cell dependent immune activation^32^. As a result, the frequencies of B cell subsets did not change significantly except reduction of mem^int^ and DN^int^ on day 21 after vaccination compared to day0 and day 7, respectively. Although not significant, a trend of increased plasmablast formation was detected at day 7 after boost (Fig. 4A). Of particular note, frequencies of mem^low^ B cells correlated strikingly with PB on day 21 and 7 days after boost. DN^low^ B cells also showed a correlation with PB 7 days after boost (Fig. 4B). These findings supported further that mem^low^ and DN^low^ expansion follows kinetics of PB induction and depends on T cell instruction as evidenced by BNT162b2 vaccination.

**Figure 4:**
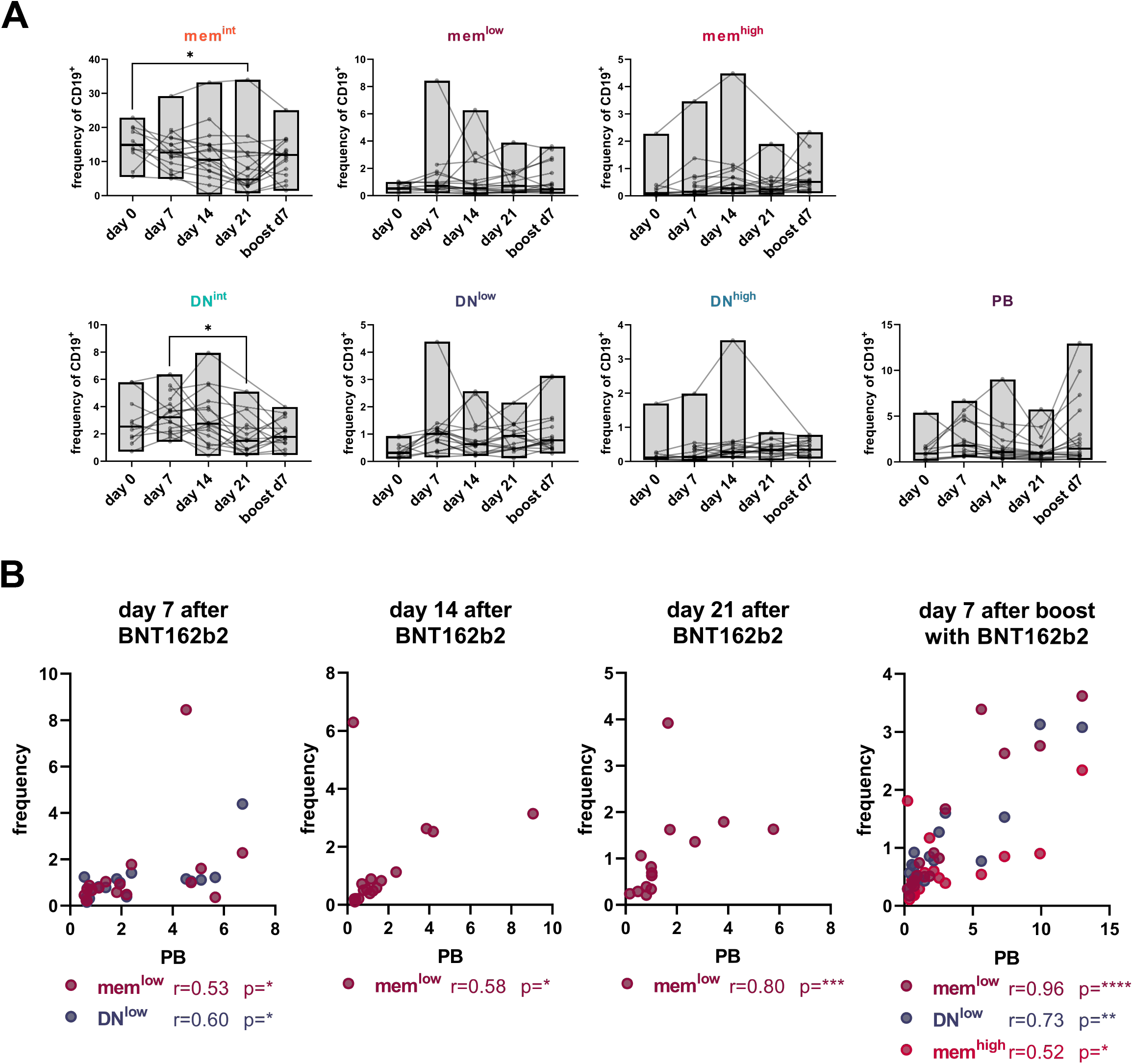
CD19^low^ (mem/DN^low^) subsets correlate with plasmablasts upon vaccination of healthy individuals with BNT162b2. **(A)** Kinetics of B cell subset and PB frequencies of 17 HDs before and 7, 14 and 21 days after first vaccination and 7 days upon boost with BNT162b2 were analyzed using flow cytometry. (KWT:\**p* ≤ 0.05, \*\**p* ≤ 0.01, \*\*\**p* ≤ 0.001, \*\*\*\**p* ≤ 0.0001) **(B)** Scatter plots show correlations between certain B cell subsets and PBs frequencies upon BNT162b2 vaccination at certain time points of HDs (n=17). (Spearman correlation: r= Spearman coefficient, \**p* ≤ 0.05, \*\**p* ≤ 0.01, \*\*\**p* ≤ 0.001, \*\*\*\**p* ≤ 0.0001).

### CD19^low^ subsets present with reduced BCR responsiveness

To evaluate B cell receptor responsiveness of newly identified subsets as a read-out for their functional competence, we studied phosphorylation kinetics of Syk (Y352) upon anti-BCR stimulation (Fig. 5A). We observed diminished Syk phosphorylation in the DN compartment compared to subsets of switched memory B cells. In both switched memory and DN B cells, subsets with low CD19 expression showed lowest pSyk kinetics while CD19^int^ and CD19^high^ subsets respond similarly to BCR stimulation. The BCR responsiveness was significantly reduced among mem^int^ and mem^high^ from patients with SLE five to eight minutes after stimulation. While there was an overall lower phosphorylation in B cells from SLE patients, both mem/DN^low^ subsets from SLE patients showed a BCR response kinetics as found for CD27^++^CD38^++^ plasmablasts (Fig. 5A).

**Figure 5:**
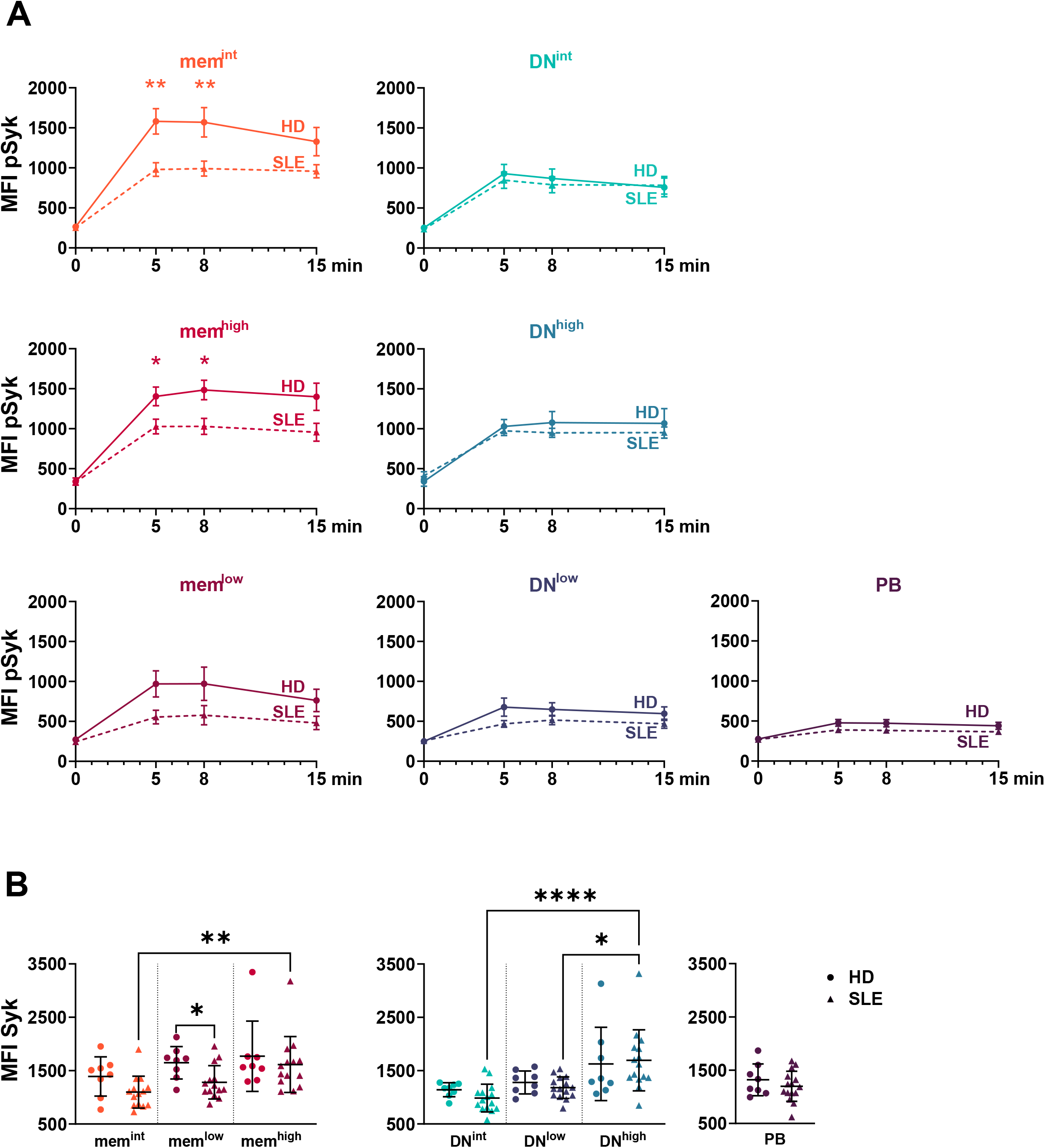
Reduced BCR responsiveness in CD19 low subsets. **(A)** Phosphorylation kinetics of Syk (Y352) of mem/DN^int^, mem/DN^low^ and mem/DN^high^ subsets and PB at time points 0, 5, 8 and 15 min in HD (n=8, solid line) and SLE (n=15, dashed line). Data is shown in means ± standard error of mean (SEM) (test: MWU between HD as SLE for each timepoint) **(B)** Scatter plot of Syk expression of mem/DN^int^, mem/DN^low^ and mem/DN^high^ subsets of HD (n=8, dots) and patients with SLE (n=15, triangle) (test: KWT).

Subsequently, we evaluated whether differences in Syk protein levels in steady state may account for this difference between subsets and SLE vs HD (Fig. 5B). We found that basal Syk levels were highest in mem/DN^high^. SLE patients showed significantly decreased Syk levels in mem^low^ B cells. Interestingly, Syk expression was comparable in mem/DN^low^ subsets compared with mem/DN^int^ cells (Fig. 5A,B).

### *PRDM1, XBP1, IRF4* and *EZH2* upregulation in CD19^low^ B cells suggests a plasmablast-like transcriptional program

To further understand the distinct nature of the analyzed B cell subsets, transcriptome analysis was performed on naive, pre-switched, total memory (IgD^-^ CD27^+^), DN^int^, DN^high^, DN^low^ and PBs of patients with SLE and HD (Fig. 6). Transcripts of IRF4 a transcription factor crucial for differentiation^33^ and survival^34^ of plasmablast and plasma cell was not only upregulated in plasmablast but also increased in DN^low^ cells. IRF4 is known to regulate Blimp-1 (encoded by *PRDM1*) expression a regulator of plasma cell differentiation^35^. The median transcription level of *PRDM1* was found increased in DN^low^ cells. Consistently, mRNA levels of *Pax5*, a transcription factor downregulated by Blimp-1, was found intermediately decreased, while *XBP1* and *EZH2* showed a slightly higher expression level among DN^low^ B cells. No changes occurred for *BCL2L1* expression as a marker for GC differentiation^36^. Thus, there was an overall trend of key transcription factors defining B cells and PB indicating that the DN^low^ population carries the transcriptional program closely related to PB.

**Figure 6:**
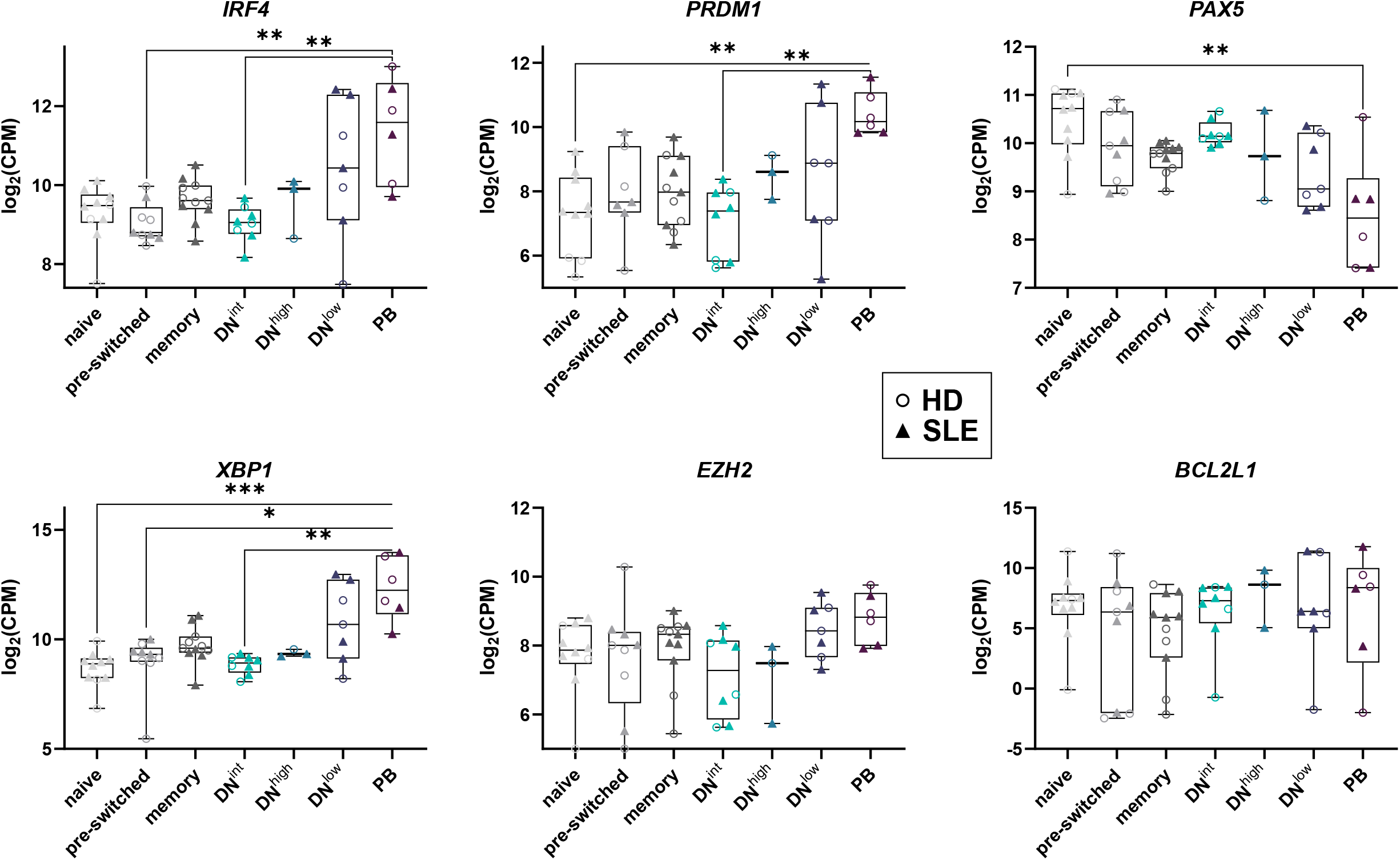
DN^low^ subsets show a transcriptional expression profile somewhat similar to plasmablasts. Boxplots of Log_2_(Counts per million(CPM)) of transcription factors *IRF4, PRDM1, PAX5, XBP1, EZH2* and *BCL2L1* depicted for naïve, pre-switched, total memory, PBs and DN subsets (DN^int^, DN^low^ and DN^high^) of each 7 donors of HD and SLE (test: KWT, *p ≤ 0.05, **p ≤ 0.01, ***p ≤ 0.001, ****p ≤ 0.0001).

## Discussion

Herein, we identified two novel CXCR5^-^CD19^low^ populations, mem^low^ and DN^low^, residing within conventional switched memory and IgD^-^CD27^-^ atypical memory B cells, respectively. Additionally, we found a not yet described CXCR5^-^CD19^high^ population in switched memory B cells (mem^high^) which shared characteristics, such as CD11c^+^ expression, with the previously reported DN2^15^. The DN2 population corresponds to the DN^high^ population which is further characterized here and has been described as precursor of antibody secreting plasmablasts generated by extrafollicular activation, and found to be increased increased in autoimmune conditions such as SLE^15^ but also acute viral infections such as SARS-CoV-2^37^. The expression profile of CD19^high^, CD38^-^, CD95^+^ and Ki-67^-^ together with the high BCR responsiveness and increased Syk expression also suggest that mem/DN^high^ represent the previously described Syk^high^ population^14^.

Although various groups reported an overall reduction of CD19 expression on B cells in SLE, a specific CD19^low^ population has not been characterized so far. In the study by Culton *et al*.^38^, the SLE cohort was subdivided into CD19^lo^ and CD19^hi^ patients based on global CD19 expression and the presence or lack of a CD19^hi^ B cell population. Autoantibodies were detected in both patient groups. The majority of the CD19^low^ B cells were described as IgD^+^, CD38^+^, and CD27^−^. Additionally, a decrease in CD19 expression on B cells was reported in anti-neutrophil cytoplasmic autoantibody associated small vessel vasculitis (ANCA-SVV) patients, suggesting that downregulation of CD19 might be a common feature of antibody driven disease^38^. Other studies reported lower CD19 expression in CD27^-^ but also in CD27^+^ B cells^39, 40^. These studies did not discriminate populations based on their IgD expression. Overall decrease of CD19 expression was seen in both active^39^ and quiescent^40^ patients with SLE and also in patients with ANCA-SVV, suggesting variation in CD19 expression as an intrinsic abnormality linked to autoimmunity rather than driven by antigen specificity or disease severity.

We found that CD19^low^ B cell subsets, mem^low^ and DN^low^ expressed co-stimulatory molecule and activation marker CD86, proliferation marker CD71^41^ and the majority were CD38^+^ and CD95^+^ while early B cell stage marker CD24^42^ and CD10^43^ were absent. In combination with surface expression of class switched immunoglobulins IgG and IgA allows the conclusion that mem/DN^low^ B cells were antigen experienced. Although, Syk itself was not reduced, phosphorylation kinetics of Syk upon anti-BCR stimulation were lower in DN^low^, similarly to PB. This could be caused either by an anergic post-activation phenotype similar to the one seen in general naïve and memory B cells of patients with autoimmune conditions like SLE, RA and pSS ^18^. An alternative explanation would be the downregulation of the BCR including BCR-associated surface molecules like negative regulator BTLA which we found downregulated in mem/DN^low^ cells. Increased frequencies of the BTLA low expressing DN^low^ population in SLE could explain the reduced BTLA expression as recently reported for the overall DN population in patients with SLE^28^.

Interestingly and in alignment with our findings, Ruschil *et al*.^44^ recently found that transcripts from total DN cells did not cluster separately from the other cell populations but instead clustered donor dependent with naïve, memory or plasmablasts. They found the number of identified differentially expressed genes was low between plasmablasts and total DN B cells^44^. In this context, our study detected that mem/DN^low^ populations clearly correlated with PBs in SLE and immune challenged HD upon vaccination with BNT162b2. In addition, targeted RNAseq analysis demonstrated an upregulated plasmablast-like transcriptional programming in DN^low^. Although further studies are needed to evaluate those findings for the mem^low^ population, the current data support that mem/DN^low^ cells are a unique subset carrying characteristics of PB destiny.

The fact that we found similar subsets in both the switched memory and DN B cell compartment was unexpected. While the double negative population is known for its heterogeneity and two of the three populations are part of the DN domain, we found shared characteristics with previously reported subsets^14, 15^. Of note, not so much is known about the diversity of the CD27^+^ memory compartment beyond Ig isotype distribution.

The features of the three subsets were comparable between corresponding subset in IgD^-^CD27^+^ switched and IgD^-^CD27^-^ atypical memory B cells supporting the idea that the increase of DN B cells in chronic immune conditions can also be largely related to loss of CD27 expression^45^ which is supported by their comparable increase in SLE and in particular by lack of CXCR5 expression. The latter can result from their a) post-GC b) extrafollicular or c) activation status also known to be related with CD27 shedding including increased solubleCD27 in autoimmune diseases^45, 46^.

It is widely known that B cell homeostasis is altered in patients with SLE and B cell targeted interventions are promising in this disease. However, the heterogeneity of the switched memory and DN populations is only partially understood, and it remains unclear how it is induced and/or maintained or how it contributes to the course of the disease. Using CD19 and CXCR5 clearly allows the differentiation of switched memory and DN B cells not only into three distinct subsets each but also suggest that mem/DN^low^ are direct precursors of plasmablasts, while mem^int^ and DN^int^ appear to belong to the B memory compartment. With new compounds targeting B cells in the pharmaceutical pipeline, it is important to understand the mechanisms of how B cells subsets are driving the disease and require consideration by innovative therapies. Our data suggest that the CXCR5^-^ populations might not be targeted by anti-CXCR5 and anti-CD19 strategies but might benefit from anti-CD38 and anti-BAFF therapies. For belimumab, which targets early, transitional B cells and partially PB/PC^47, 48^, recent studies could document that blocking BAFF/BLyS by belimumab had rapid effects on B cell subsets of earlier developmental stages such as naïve B cells. Late B cell stages, such as memory or plasma cells, decreased later in a gradual manner or did not change upon treatment. Only early immunological changes correlated with disease improvement^49^ These data together with our study provide a rationale for modalities that target naïve and early B cell stages and may thereby prevent not only their differentiation into memory B cells but also their direct path becoming plasmablasts/plasma cells.

Collectively, the data presented here, including surface marker expression, correlation analysis, BCR kinetics and transcription analysis strongly indicate that mem/DN^low^ cells are precursors of PB and directly contribute to plasmacytosis upon immune activation. These mem/DN^low^ cells reflect a subset of pre-plasma cells that may not require to undergo full or incomplete memory B cell differentiation. Understanding of the involved selection mechanisms will be important not only in terms of their immunobiological features but also for potential treatment strategies. In this regard, the current data suggest that there could be selective treatment approaches not only for certain B cell subsets but also distinct PB/PC compartments, including the possibility to leave protective PB/PC untouched.

## Data Availability

The datasets used and analyzed during the current study are available from the corresponding author on reasonable request.

## Acknowledgment

We thank S. Gaertner, D. Hurd and M. Rastegar from HTG Molecular Diagnostics, Inc. and J. Kirsch, T. Kaiser from the Flow Cytometry Core Facility of the DRFZ.

## Author Contributions

The theoretical framework was developed by FS, ACL and TD.

Data were obtained by FS, ALS, ES, HR-A, AW, KR, ML, VDD, AF and SF.

Data were analyzed by FS, ACL.

All authors developed, read, and approved of the current manuscript.

## Ethics Statement

This study was carried out in accordance with the recommendations of the ethics’ committee at the Charité University Hospital Berlin with written informed consent from all subjects. All subjects gave written informed consent in accordance with the Declaration of Helsinki.

## Figure legends

**Supplementary Figure 1: Surface expression patterns on mem**^**int**^, **mem**^**low**^ **and mem**^**high**^ **detected by flow cytometry. (A)** Gating strategy of flow cytometry staining for identification of mem and DN subsets shown for a HD. **(B)** Box and whisker plots of median FI of CD71, CD24, CD11c and frequencies of CD38^+^ or CD95^+^ of HD (n=8) for each subset. **(C)** Box and whisker plots of median FI of PD1 and PD-L1 for each subset. **(D)** Box and whisker plots of median FI of CD71 and frequencies of CD95^+^ and CD38^+^ cells memory subsets for HD (dots, n=8) compared to SLE patients (triangles, n=9). (Significance levels: *p ≤ 0.05, **p ≤ 0.01, ***p ≤ 0.001, ****p ≤ 0.0001).

**Supplementary Figure 2: Activation marker and checkpoint molecule expression detected by mass cytometry (A)** Gating strategy of CyTOF. **(B)** Distribution of subsets within IgD^-^CD27^+^ and IgD^-^CD27^-^ **(C)** Scatter plots show expression of CD45RA and CD45RO for comparison in between subsets.**(D)** CD45RA and CD45RO differences between HD and patients with SLE. Scatter plots show expression of PD1 and PD-L1 for comparison in between **(E)** subsets or **(F)** between HD (dots) and SLE patients (triangles). **(G)** Scatter plots show expression of Ki-67 for comparison in between subsets or between HD (dots) and SLE patients (triangles). (Data is shown as median + 95%CI of [n(HD/SLE) = 18/24]. (Significance levels: *p ≤ 0.05, **p ≤ 0.01, ***p ≤ 0.001, ****p ≤ 0.0001)

**Supplementary table1:**
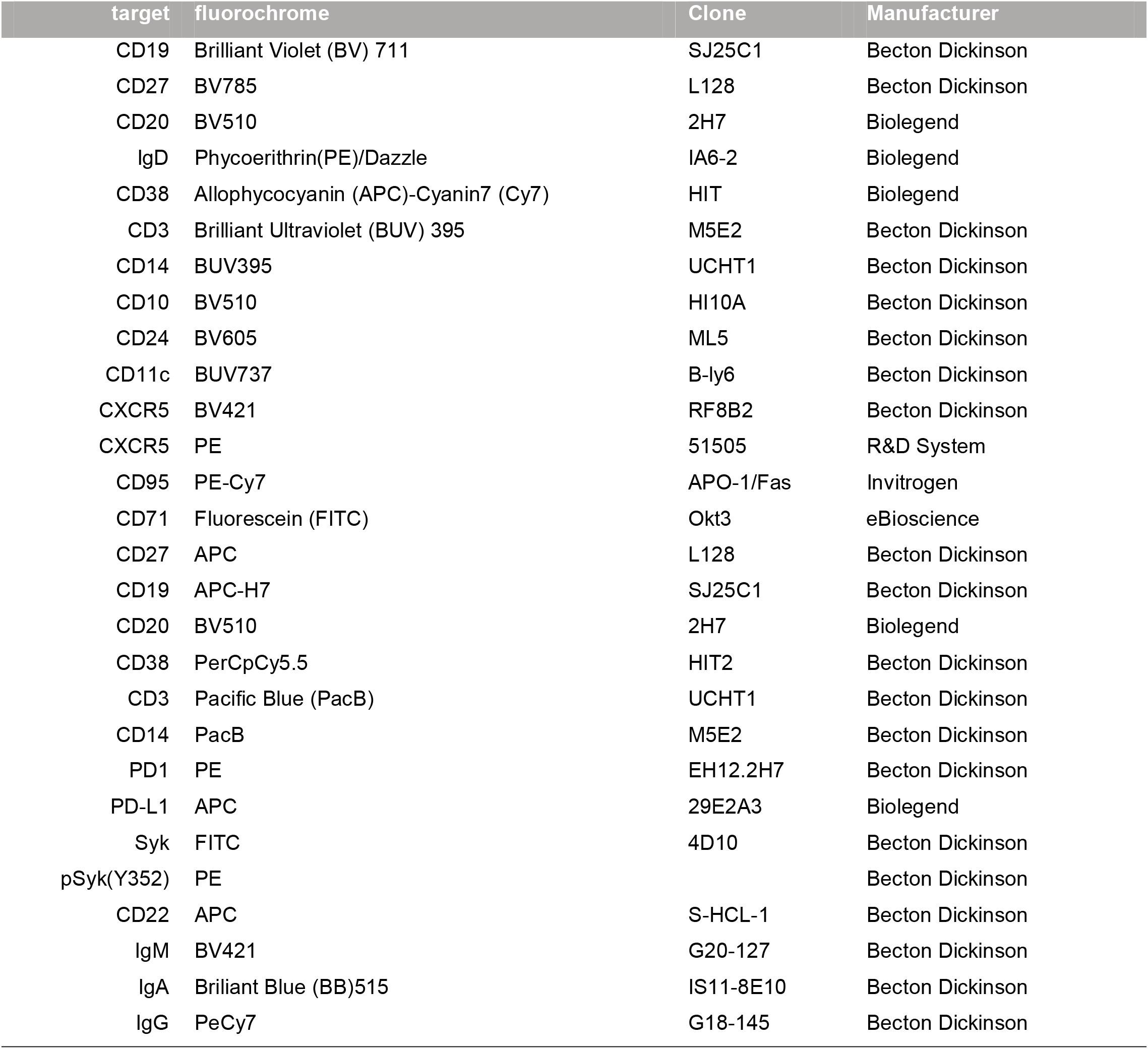
Antibodies for flow cytometry analysis.

**Supplementary table 2:** Antibodies for mass cytometry analysis.

| antibody | Isotype tag |
| --- | --- |
| CD45 | Y89 |
| ICOS | Pr141 |
| CD19 | Nd142 |
| VISTA | 908-Nd143 |
| CD69 | Nd144 |
| CD4 | Nd145 |
| IgD | Nd146 |
| CD11c | Sm147 |
| CD274/PD-L1 | Nd148 |
| CD25 | Sm149 |
| CD223/LAG-3 | Nd150 |
| CD123 | Eu151 |
| TCRgd | Sm152 |
| TIGIT | Eu153 |
| TIM3 | Sm154 |
| CD27 | Gd155 |
| CD86/B7.2 | Gd156 |
| CD137/4-1BB | Gd158 |
| Foxp3 | Tb159 |
| CD14 | Gd160 |
| CD152/CTLA-4 | Dy161 |
| CD8 | Dy162 |
| CD272/BTLA | Dy163 |
| CXCR5 | Dy164 |
| CD45R0 | Ho165 |
| CD155 | Er166 |
| CD38 | Er167 |
| Ki-67 | Er168 |
| CD45RA | Tm169 |
| CD3 | Er170 |
| CD226 | Yb171 |
| IgM | Yb172 |
| CD162/PSGL | Yb173 |
| HLA-DR | Yb174 |
| CD279/PD-1 | Lu175 |
| CD56 | Yb176 |
| CD16 | Bi209 |

