## Supplemental Material for "Antigen-experienced CXCR5^-^ CD19^low^ B cells are plasmablast precursors expanded in SLE"

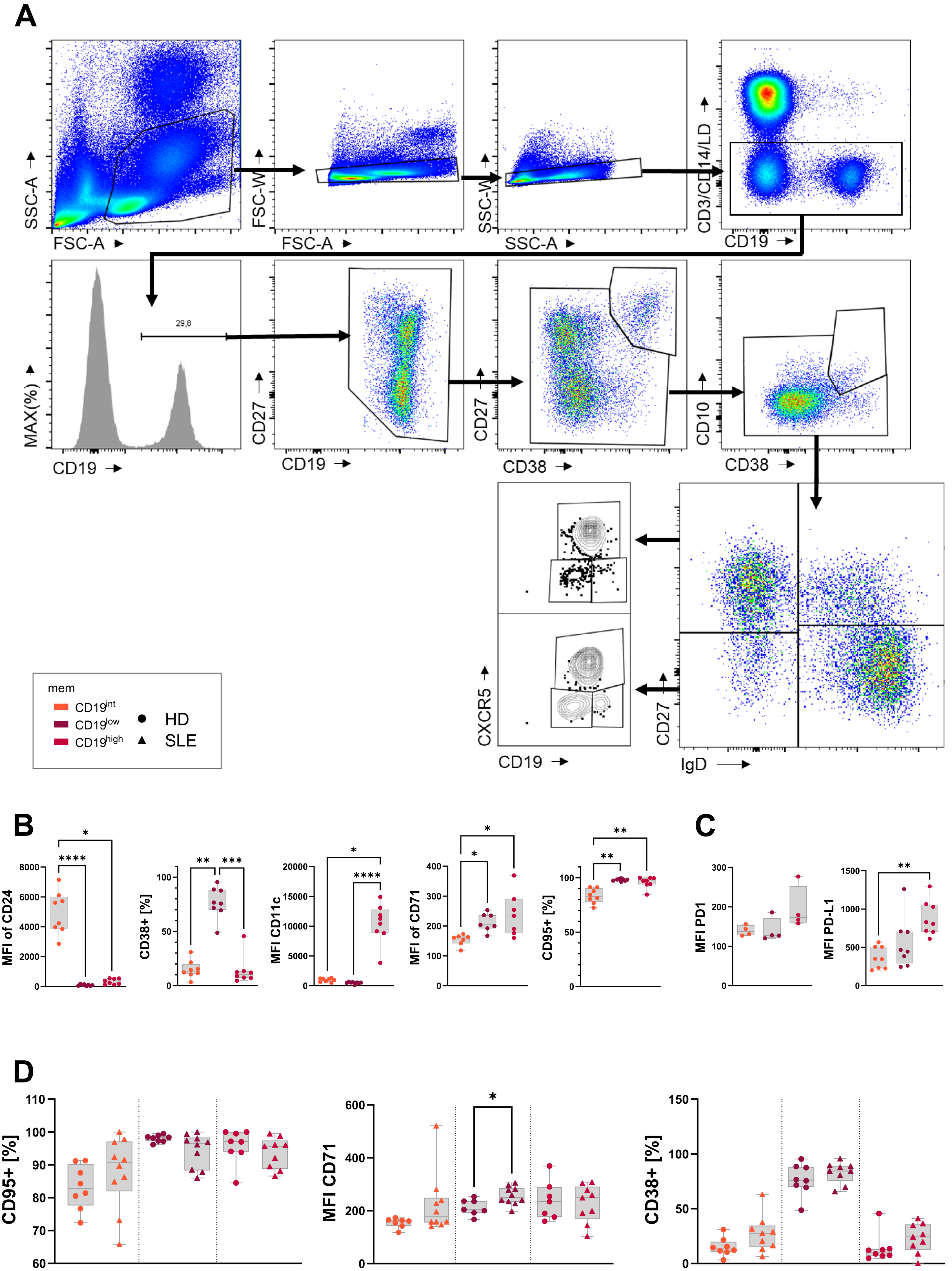


Supplementary Figure 1
Szelinski et al.

**Supplementary Figure 1: Surface expression patterns on mem$^{int}$, mem$^{low}$ and mem$^{high}$ detected by flow cytometry. (A)** Gating strategy of flow cytometry staining for identification of mem and DN subsets shown for a HD. **(B)** Box and whisker plots of median FI of CD71, CD24, CD11c and frequencies of CD38$^{+}$ or CD95$^{+}$ of HD (n=8) for each subset. **(C)** Box and whisker plots of median FI of PD1 and PD-L1 for each subset. **(D)** Box and whisker plots of median FI of CD71 and frequencies of CD95$^{+}$ and CD38$^{+}$ cells memory subsets for HD (dots, n=8) compared to SLE patients (triangles, n=9). (Significance levels: *p ≤ 0.05, **p ≤ 0.01, ***p ≤ 0.001, ****p ≤ 0.0001).

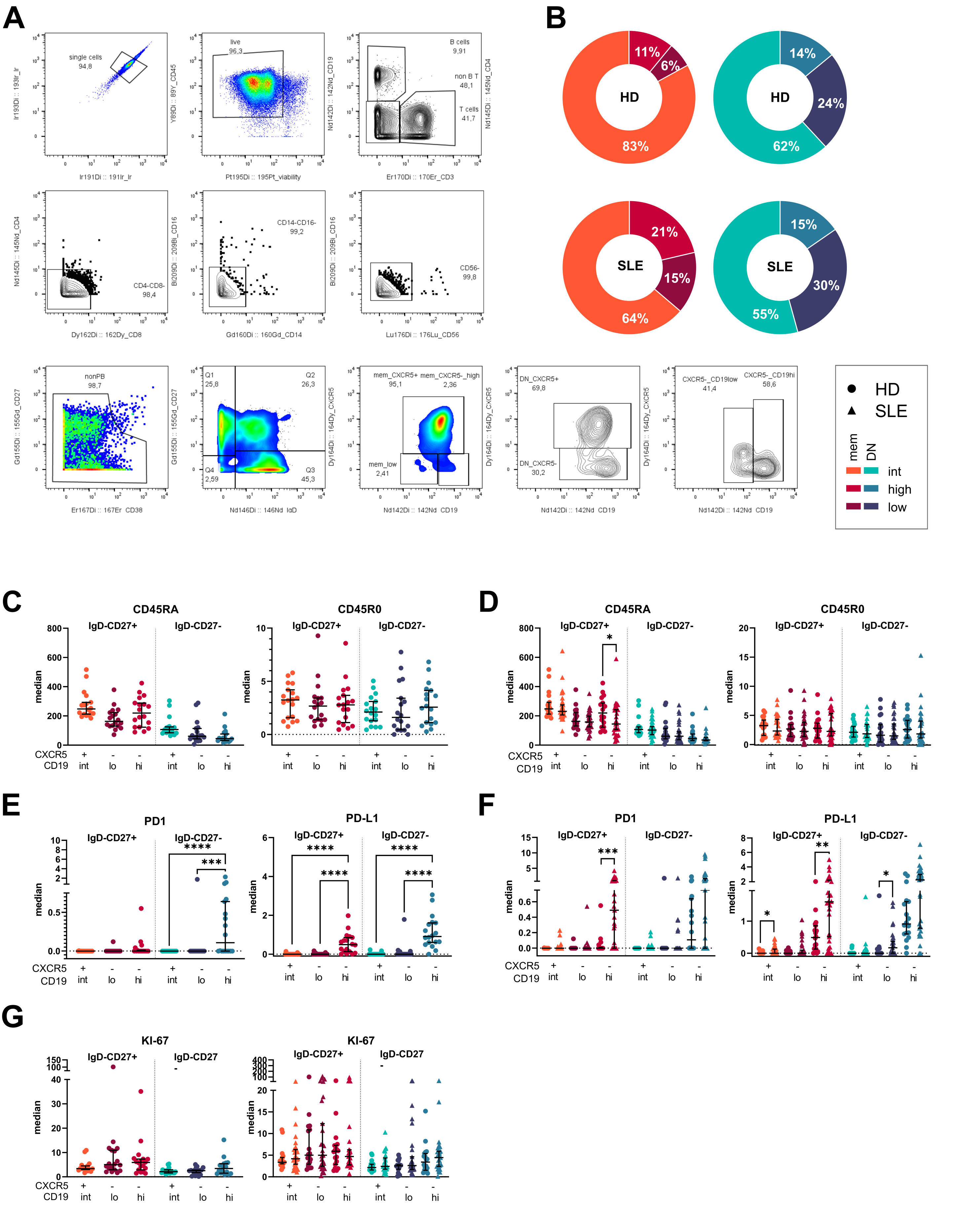


Supplementary Figure 2
Szelinski et al.

**Supplementary Figure 2: Activation marker and checkpoint molecule expression detected by mass cytometry (A)** Gating strategy of CyTOF. **(B)** Distribution of subsets within $IgD^{-}CD27^{+}$ and $IgD^{-}CD27^{-}$ **(C)** Scatter plots show expression of CD45RA and CD45RO for comparison in between subsets.**(D)** CD45RA and CD45RO differences between HD and patients with SLE. Scatter plots show expression of PD1 and PD-L1 for comparison in between **(E)** subsets or **(F)** between HD (dots) and SLE patients (triangles). **(G)** Scatter plots show expression of Ki-67 for comparison in between subsets or between HD (dots) and SLE patients (triangles). (Data is shown as median + 95%CI of [n(HD/SLE) = 18/24]. (Significance levels: *$p \leq 0.05$, **$p \leq 0.01$, ***$p \leq 0.001$, ****$p \leq 0.0001$)
